# Genome Wide Association Analysis of Two Pore Domain Potassium Channel Gene Regions Reveals Multiple Pain-Associated Signals

**DOI:** 10.64898/2025.12.24.25342351

**Authors:** Samuel R Bourne, Jenny Cook, Paul D Wright, Jeffrey Jerman, Rachel Forfar, Emma L Veale, Alistair Mathie

## Abstract

Two-pore domain potassium (K2P) channels are novel analgesic drug targets with existing *in vitro* and *in vivo* validation, however genetic associations in large scale populations have not been widely reported. By examining two comprehensive genetic databases (UK Biobank & FinnGen) and 136 pain-related traits, we highlight that multiple pain-associated signals are present within 500 kb of K2P gene transcripts, with particularly strong evidence for KCNK5 (TASK-2) and headache-related traits. This data benchmarks remarkably well against gene targets already considered to be well validated pain targets (voltage-gated sodium and transient receptor potential channels) and is enhanced when examining populations regularly consuming analgesics who also report pain. This data supports a rationale for developing therapeutic strategies to modulate K2P function in patient populations experiencing pain.

## Introduction

Chronic pain, often defined as a pain persisting for longer than three months, affects around 20-25% of the adult population worldwide and represents a major public health burden. Conditions such as low back pain, neck pain, migraine, arthritis, and other musculoskeletal pain conditions are among the most prevalent causes of chronic pain and leading contributors to long-term disability (Vos et al., 2020, Zhu et al., 2024). Despite the increasing prevalence of chronic pain, there remains a clear and urgent need for novel, efficacious therapeutic strategies. However, there are multiple interrelated challenges that significantly hinder therapeutic development in this disease. These include the marked heterogeneity of the chronic pain patient population, both in terms of clinical presentation and underlying pathophysiology (Vellucci, 2012); the polygenic and multifactorial nature of the disease pathogenesis (Mogil, 2012); and the limited translatability of current animal models, which often fail to replicate the human pain phenotype (Burma et al., 2016). Additionally, a pronounced and variable placebo effect complicates the interpretation of clinical trial outcomes (Cragg et al., 2016), while the absence of robust and reliable clinical biomarkers continues to impede diagnosis, stratification, and treatment monitoring (Mackey et al., 2025). Collectively, these factors contribute to the high attrition rate observed in chronic pain drug development and limit the successful translation of promising preclinical targets into clinically approved treatments.

To address these barriers there has been a strategic shift towards leveraging human genetic data derived from large patient data biobanks and genome-wide association studies (GWAS). There is growing evidence to show that drug targets supported by human genetic associations, including rare coding loss- or gain-of-function mutations or common genetic variants are almost twice as likely to succeed in clinical development, compared to those without genetic insights (Nelson et al., 2015, Ochoa et al., 2022). A number of recent GWAS studies have identified numerous common genetic variants associated with chronic pain phenotypes (Pan et al., 2025, Johnston et al., 2019, Suri et al., 2018).

Perhaps unsurprisingly, given their central role in regulating action potential initiation and propagation, and their high expression in nociceptive neurons, ion channels have emerged as key contributors to pain susceptibility and persistence. GWAS studies have identified variants in genes encoding several ion channels, include the voltage-gated sodium channels (SCN9A, SCN10A), sodium-activated potassium channel (KCNT2), calcium activated potassium channel (KCNN2) and transient receptor potential (TRP) channels (Williams et al., 2020, Akerlund et al., 2024, Packer et al., 2025). With a burgeoning, global, opioid epidemic, the selective targeting of peripheral ion channels has emerged as a promising non-opioid strategy to treat chronic pain.

Most notably, the recent FDA approval of suzetrigine (JOURNAVX, VX-548), a first-in-class peripherally selective Nav1.8 sodium channel blocker, for the treatment of moderate to severe acute pain, marks a pivotal moment in ion channel-based analgesia (Bertoch et al., 2025). This approval marks a paradigm shift towards precision ion-channel modulation, rather than broad CNS suppression using opioids.

Despite extensive functional and experimental evidence supporting the role of two-pore domain potassium (K2P) channels, particularly, KCNK2 (TREK1), KCNK4 (TRAAK) and KCNK18 (TRESK), in nociceptive signalling and inflammation (Alloui et al., 2006, Dobler et al.,2007, Brohawn et al., 2014), direct genetic associations with human pain phenotypes have remained limited. A rare truncating mutation in KCNK18 (TRESK), was previously identified in a small cohort of patients experiencing familial migraine with aura, providing early human genetic support for the involvement of K2P channels in pain (Lafrenière et al., 2010, Andres-Enguix et al., 2012). However, to date, no GWAS or large-scale biobank analyses, have directly associated K2P channels in chronic pain phenotypes, nor replicated associations between KCNK18 and common migraine (Markel and Curtis, 2022).

In this study we provide large-scale genetic evidence to associate variants within K2P channel gene regions with chronic pain phenotypes in data from the Pan-UK Biobank and FinnGen cohorts. Highlighting KCNK5 (TASK2) and KCNK10 (TREK2) specifically as compelling candidates within these datasets. These finding establish a genetic basis for K2P channel involvement in human pain and highlight this family of ion channels as promising therapeutic targets for novel analgesics.

## Results

### 38 variants within ±500 kb of K2P genes are strongly associated with pain-related traits and another 205 are suggestively linked

The total pool of assessed variants included approximately 40% common (minor allele frequency > 0.1%), 42% rare (> 0.01%) and 18% ultra-rare (< 0.01%) single nucleotide polymorphisms (SNPs). Out of 136,166 Pan-UK Biobank (BB) and 100,662 FinnGen variants within ±500 kb of K2P gene transcripts tested for association with 136 pain-related traits, 38 surpassed the Bonferroni-corrected significance threshold (-Log P-value >6.3, P<0.5×10^6^) – *Figure 1, Table 1 & supplementary table 3*, and 205 met the threshold for suggestive significance (-Log P >5.3, P<0.5×10^5^) - *supplementary table 4*. The strongest associations included analgesic medication use (e.g. pregabalin prescriptions, paracetamol prescriptions and regular ibuprofen use), headache, chest pain, neuropathies, and undergoing treatment with a pain consultant, and were distributed across 8 of the 13 K2P gene regions (KCNK1, KCNK3, KCNK5/16/17, KCNK7, KCNK9, KCNK10, KCNK13 and KCNK18). While the pool of suggestive SNPs (n=205) also alluded to a wider general contribution of these gene regions to pain (*supplementary table 4)*.

**Figure 1.**
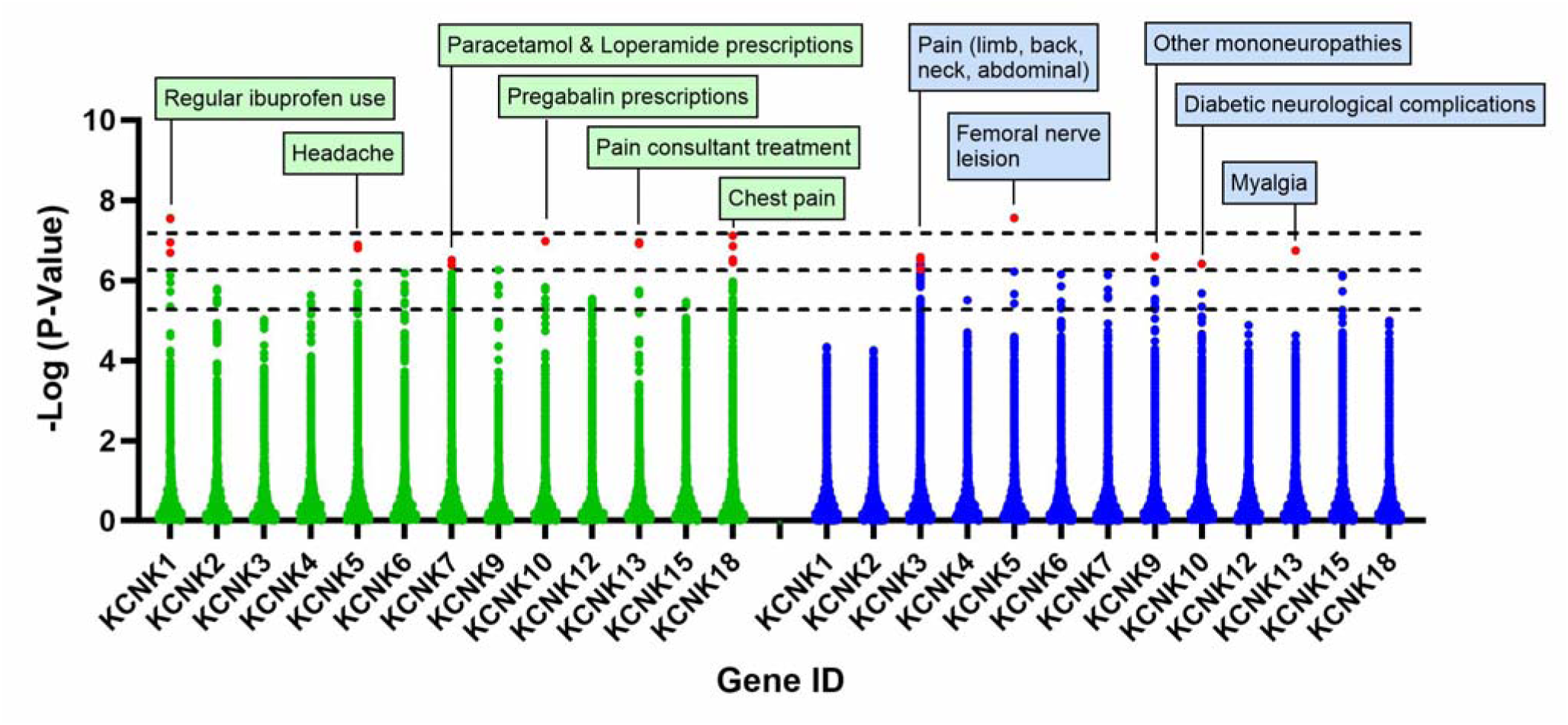
Association of 136,166 Pan-UK Biobank (green) and 100,622 FinnGen (blue) variants located within ±500 kb of K2P genes with 136 pain-related traits. Significance thresholds are indicated by horizontal dashed lines: suggestive (-Log P > 5.3), Bonferroni-corrected (-Log P > 6.3) and global (-Log P > 7.3). Variants surpassing the Bonferonni threshold are depicted in red and labelled with the associated trait.

**Table 1.**
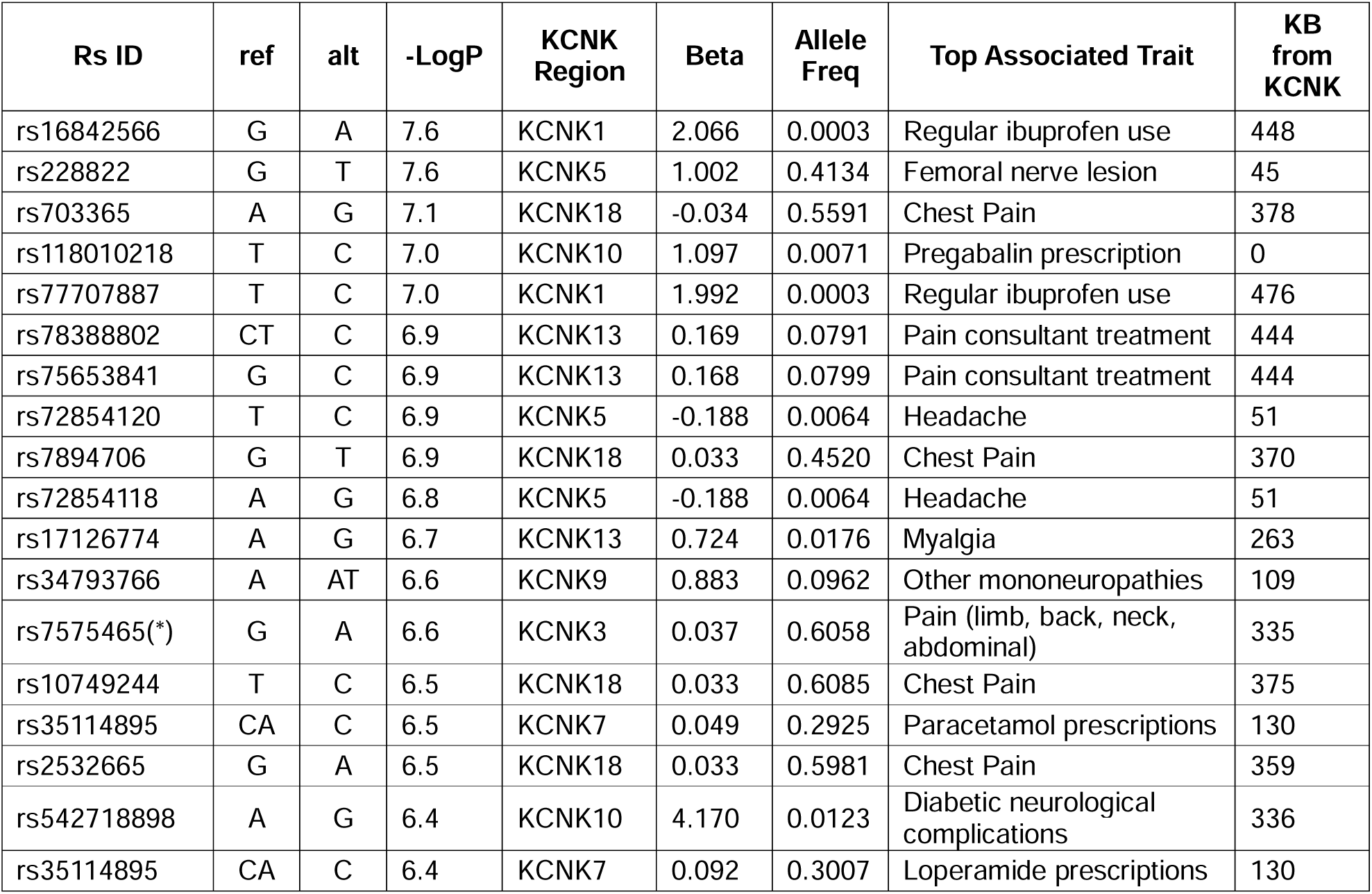
Variants significantly associated with pain-related traits (Bonferonni threshold, -Log P > 6.3) in the Pan-UK BB and FinnGen datasets, located within ±500 kb of K2P (KCNK) genes. Columns include variant ID (Rs ID), reference allele (ref), alterative allele (alt), -log(p-value) (-LogP), effect size (beta), allele frequency (allele freq), associated phenotype (top associated trait), and distance from the nearest KCNK gene in kilobases (kb from KCNK). N.B. The KCNK5 region also includes variants in close proximity (<100 kb) to KCNK16 and KCNK17. (*) One of 22-linked variants shown for KCNK3, clustered in the same region and in association with the same trait. See supplementary table 3 for full list and details.

It should be noted that due to the close genomic proximity of KCNK5, KCNK16, and KCNK17 (within <100 kb), associations in these regions (±500 kb) were underpinned by the same variant(s). Interestingly, 22 of the Bonferroni-corrected significant variants were clustered within the same location of the KCNK3 gene region (±500 kb), suggesting these may represent a single pain-associated signal, supported by multiple linked variants. However, this cluster was 300-400 kb from the KCNK3 gene itself reducing confidence a resulting phenotype is driven by this transcript specifically.

#### Detection of pain-linked signals in proximity of K2P gene transcripts benchmarks well against voltage-gated sodium channel and transient receptor ion channel gene regions, which are considered validated targets for pain

To contextualize and benchmark the pain-associated signals detected in K2P gene regions, we applied the same analytical approach to all variants within ±500 KB of voltage-gated sodium channel (SCNA) and transient receptor potential (TRPV) channel genes, evaluating their association with the 87 pain-related traits from the Pan-UK BB (figure *2*). Interestingly, not only was the strength of K2P gene associations comparable with those in SCNA and TRPV gene regions, but in some cases surpassed them. Notably, a greater diversity of pain phenotypes reached Bonferroni-corrected significance in K2P gene loci compared to SCNA and TRPV regions. Furthermore 6 out of 13 K2P gene regions harboured at least one variant surpassing the Bonferonni threshold for significance (-log P > 6.3), compared to 5 out of 9 SCNA and 3 out of 6 TRPV regions (*supplementary figure 1*). There was also evidence of gene region-specificity with respect to the types of pain traits associated. For example, headache was the only trait which shared significant associations across all the three gene families (KCNK5/16/17, SCN3A, SCN2A and TRPV4). In contrast, back pain was specifically linked to SCN5A, SCN10A and SCN11A, while gabapentin prescriptions were significantly associated with TRPV5 and TRPV6 regions. K2P regions, however, exhibited associations with a wider assortment of pain-related traits, which could reflect a more diverse functional involvement.

**Figure 2.**
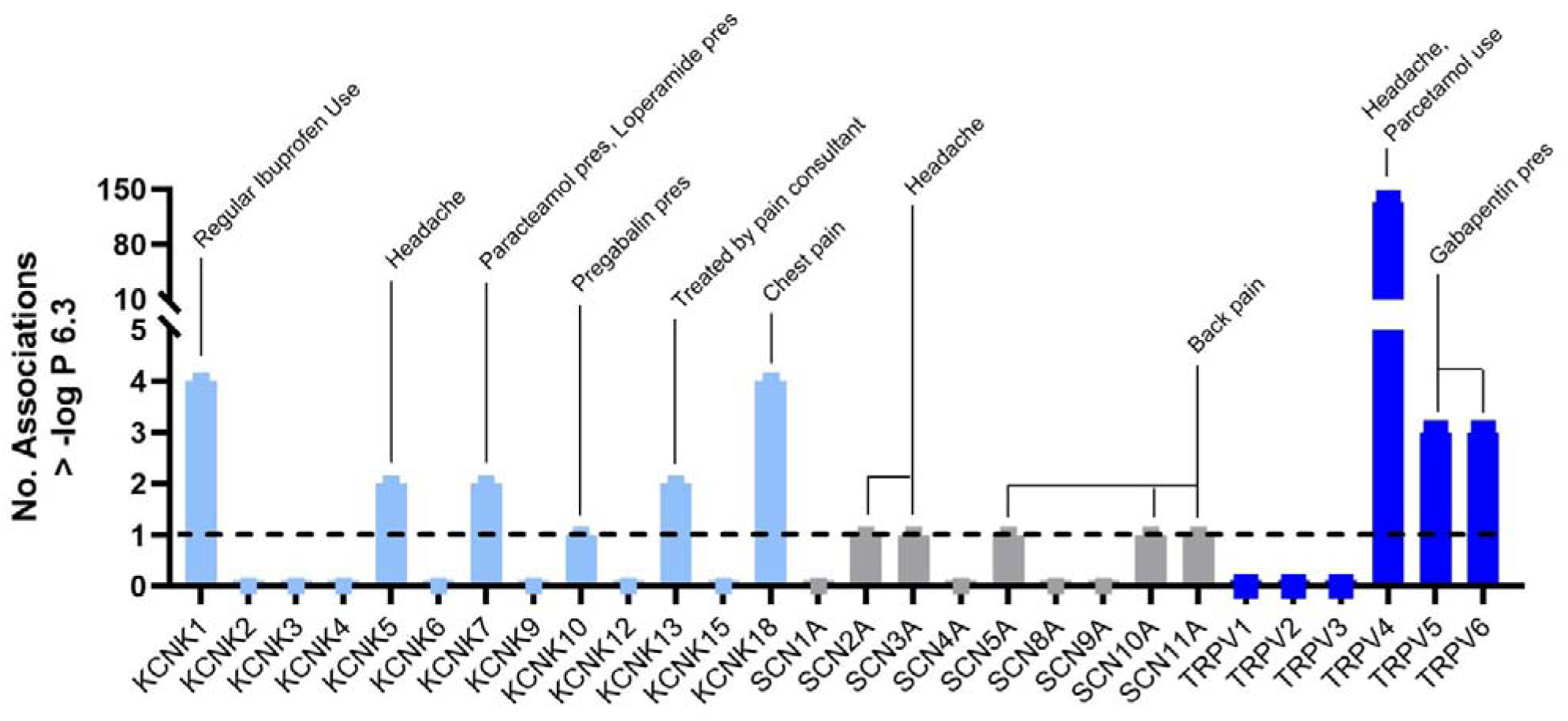
Diversity of pain conditions linked to K2P, SCNA and TRPV gene regions ±500 kb. Comparison of the number of Bonferroni-corrected variants (-Log P > 6.3) within ±500 kb of K2P, SCNA and TRPV gene transcripts linked with pain-related traits in the Pan-UK BB with the top associated traits labelled.

Additionally, the same methodology was applied to randomly selected, well-powered alternative traits to explore the specificity of K2P gene region signals to pain. Importantly, no significant associations were identified between variants within ±500 kb of K2P genes and asthma, ischemic heart disease or peripheral nerve disorders and only one was found for mental disorders (*supplementary figure 2*). This supports trait specificity for the relationship between K2P variants and pain phenotypes.

### Mapping of pain-linked variants within K2P gene regions suggests possible effects on gene expression and regulation driven from non-coding regions

All pain-associated variants were located in non-coding regions, with the majority mapping to intergenic (between genes) or intronic (between exons) DNA. Of those 38 variants that surpassed Bonferroni-corrected significance, two overlapped with open chromatin regions, four were located in promoter flanking regions, six in promoters and one within CpG islands (*supplementary table 3*). Only one variant mapped within a K2P transcript itself, an intronic variant in KCNK10 (rs118010218) which was significantly associated with pregabalin prescriptions. 30 of those highly significant variants also scored strongly for likelihood to be functional (Regulome DB score > 0.5) (*Supplementary table 5*). Although, most of these variants occurred more than 100 kb from K2P gene transcripts, two, rs72854120 and rs72854118 were within a regulatory region 50 kb upstream of KCNK5 and downstream of KCNK16/17, respectively. These variants showed strong association with headache (-LogP > 6.3) alongside variants downstream of KCNK5 which were suggestively linked (-LogP > 5.3) to regular paracetamol use (*supplementary table 4*).

Although not meeting Bonferroni-corrected significance, 65 of the 205 suggestively associated variants (-Log P > 5.3) were located within key regulatory regions including enhancers, promoters, CpG islands, and transcription factor binding sites (*supplementary table 4*). Of these, 22 variants were located within 50 kb of K2P gene transcripts (*supplementary figure 3*) Including: KCNK1 (abdominal pain), KCNK5 (paracetamol prescriptions and headache), KCNK6 (back pain), KCNK7 (paracetamol, loperamide prescriptions and general pain), KCNK10 (pregabalin prescriptions), KCNK12 (knee pain >3 months) and KCNK18 (abdominal pain).

### Linkage disequilibrium between pain-linked variants within K2P gene regions increases confidence in these trait-linked signals and reinforces evidence linking the KCNK5 region to headache

Strong LD (R^2^ > 0.5), which suggests variants are co-inherited, was observed among multiple variants associated with the same pain-related traits (*supplementary table 6*). This increases the weight of evidence for these signals within K2P gene regions, because it hints at the existence of multiple pain-related variants which are exerting effects on gene function at these loci. Indeed, of the all variants with P > -Log 5.3 (suggestive threshold), 79% were in moderate LD (R^2^ > 0.5) with one another, highlighting this. This was most compelling for the KCNK5 region because of the strength of association and proximity to the K2P transcript. Importantly, strong LD was also observed between rs72854120, rs72854118 and rs72851880, reinforcing the significance of the KCNK5 locus for headache susceptibility (figure *3*). Although this genomic region contains transcripts for several other genes including GLO1 (glyoxalase I), DNAH8 (dynein axonemal heavy chain), GLPR1 (glucagon-like peptide), SAYSD1 (SAYSvFN domain-containing protein 1), and KIF6 (kinesin family member 6), the majority of the pain-associated variants were clustered in the vicinity of KCNK5, KCNK16, and KCNK17 (figure *3.A*), highlighting the potential importance of this K2P gene cluster in the observed pain associations.

**Figure 3.**
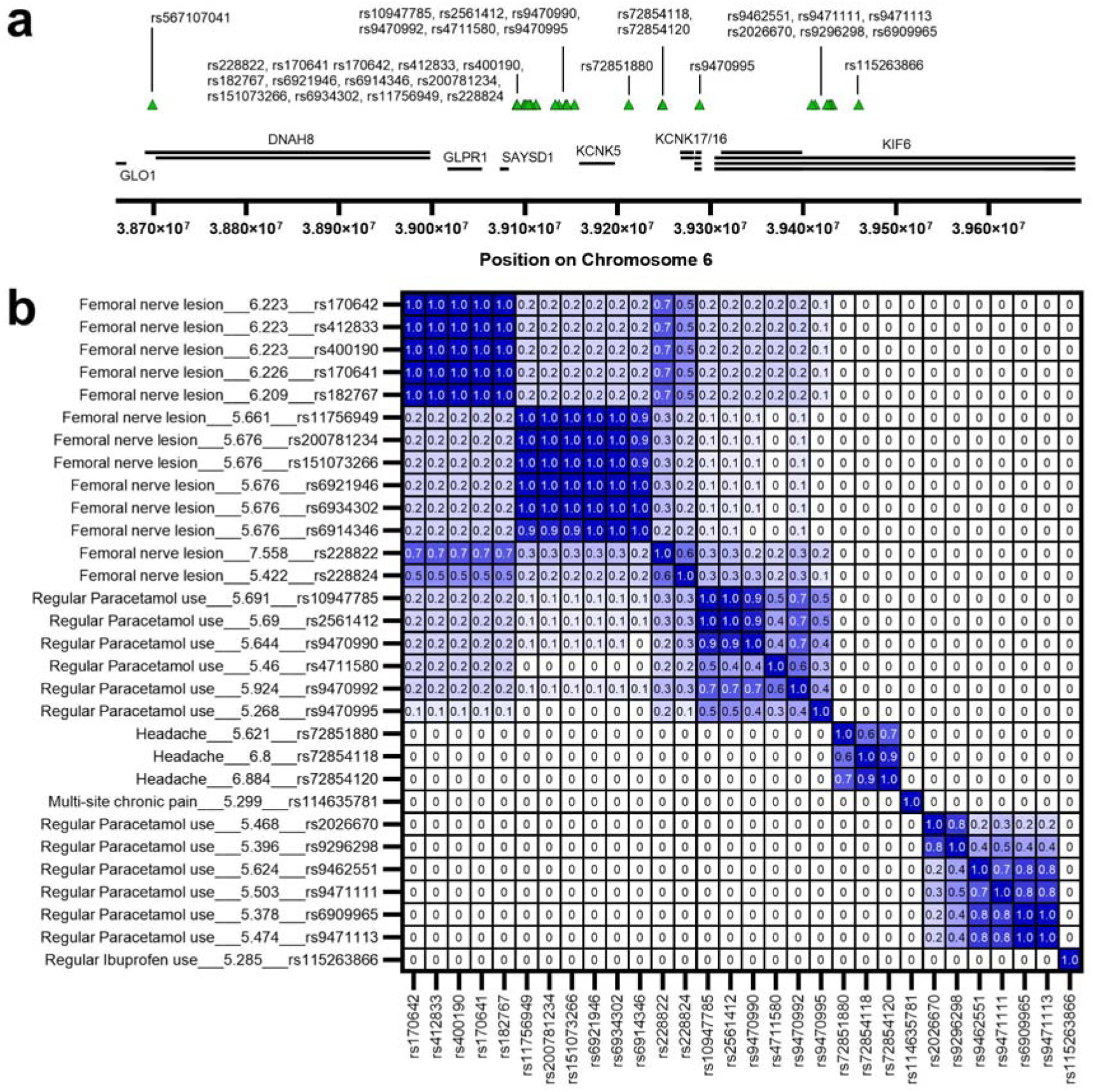
Mapping of pain-associated variants on chromosome 6, located within ±500 kb of KCNK5 (A). Variants suggestively (-Log P > 5.3) or significantly (-Log P > 6.3) associated with pain-related traits are shown as green tringles and the surrounding gene transcripts as black bars (A). Linkage disequilibrium (LD) scores of pain-associated variants on chromosome 6, located within ±500 kb of KCNK5 (B). LD is shown as R^2^ values, with each variant annotated from left to right with the associated trait, -Log P-value and RsID number.

Further examination revealed that some SNPs were in LD with other variants located within ±500 kb of the same K2P gene loci that are strongly associated with non-pain-related traits (*supplementary table 7 & 8*). These included variants in weaker LD (R^2^ > 0.2) with associations for bipolar disorder & Crohn’s disease (KCNK10), cortisone levels (KCNK12), platelet count (KCNK15), neuroticism and anxiety (KCNK18). In addition, strong LD (R^2^ > 0.8) was observed with variants linked to white blood cell counts (KCNK3), reticulocyte count (KCNK4), fasting plasma glucose and migraine (KCNK5), depression and inflammatory bowel disease (KCNK7), circulating phylloquinone levels (KCNK9) and pulmonary artery enlargement (KCNK10). In addition, four variants within the K2P gene loci themselves had previously been reported in the GWAS Catalog as genome-wide significant (-Log P > 7.3) for other traits: rs7575465 (cortical surface area and schizophrenia), rs34732613 (cortical thickness) and rs11126857 (associated with CGREF1/FAM3C protein level ratio) in the KCNK3 region and rs72854120 in the KCNK5 region (with headache or migraine) (*supplementary table 9*). To our knowledge none of the other remaining lead variants have been strongly associated with other traits. These findings suggest a broader biological relevance of K2P gene regions in both pain and other complex traits

### A multivariate analysis of pain and regular analgesic use enhances the association of gene variants within ±500 KB of K2P transcripts, with strong evidence for KCNK5 and KCNK10

To explore the effects of pain-related traits and chronic analgesic use, a multivariate analysis was performed using all common Pan-UK BB variants (allele frequency > 1%) to test for significance. Each non-medication pain-related trait was analysed pairwise in combination with either regular use of ibuprofen, paracetamol or aspirin (defined as use on most days for the last 4 weeks). A variant was considered as significantly associated if it’s multivariate p-value exceeded the Bonferroni-corrected threshold (-Log P-value > 6.3) and it showed at least one order of magnitude increase relative to the corresponding univariate association. Using this approach, six of the K2P gene regions (±500 kb) met these criteria for multivariate significance: KCNK5, KCNK7, KCNK9, KCNK10, KCNK13 and KCNK18 (*supplementary figure 10*). Among these, the KCNK5 and KCNK10 gene loci yielded variants within < 25 kb of K2P transcripts (figure *4. A&B*). These included associations between pain and regular paracetamol use in the KCNK5 (rs10947785, rs2561412, rs9470990, rs755853, *Figure 4.A*) and KCNK10 (rs4899948, *Figure 4.B*) gene regions. Notably, rs10947785, rs2561412, rs9470990 and rs755853 clustered approximately 20 kb downstream of KCNK5 in a region encompassing enhancers and were in strong LD (R^2^ > 0.8) with a nearby variant previously linked to fasting plasma glucose, as well as in weak LD (R^2^ > 0.2) with variants linked to coronary artery disease and red blood cell count (*supplementary table 7 & 8*).

**Figure 4.**
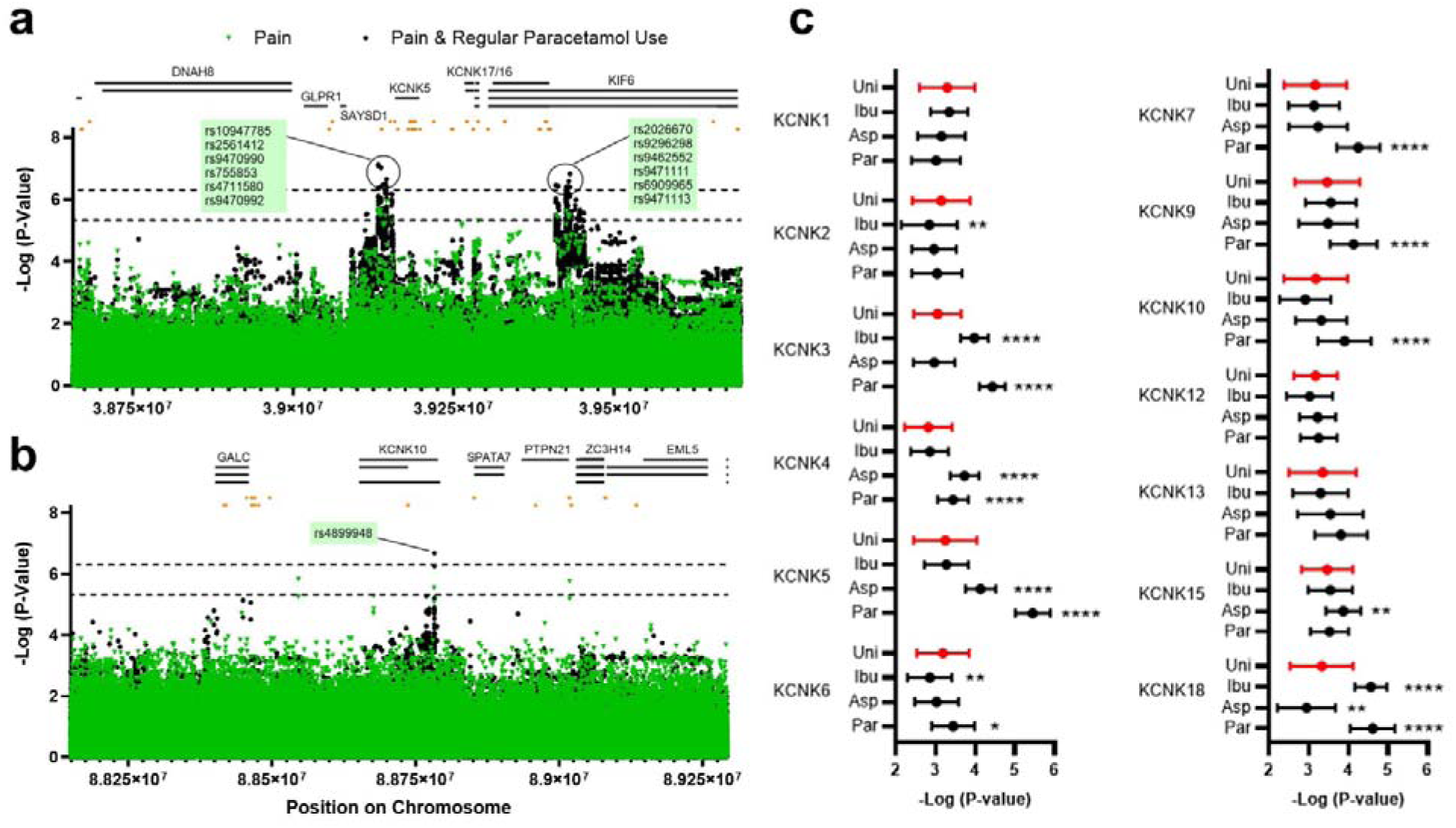
Pairwise multivariate analyses between Pan-UK BB pain-related traits and regular use of either aspirin, paracetamol, or ibuprofen for common gene variants within 500 KB of K2P gene transcripts. Mapping of associations within the KCNK5 (A) and KCNK10 (B) gene regions for pain (univariate analysis) or pain and regular paracetamol use (multivariate analysis). Gene transcripts are represented by black bars, with enhancers/promoters indicated as orange bars. The mean and standard deviation for the top association statistics (-Log P-value) for pain-related traits and K2P gene loci variants (C), distinguished by univariate analysis (uni), ibuprofen (Ibu), paracetamol (par) or aspirin (asp) multivariate analyses. Significance was tested by using a two-way ANOVA (Dunnett’s post-hoc test). Whereby P < 0.00005 (****), P < 0.0005 (***), P < 0.005 (**), P < 0.05 (*).

By compiling the top association statistic for each trait, we summarised the effects of the multivariate analysis for pain and regular analgesic use, comparing these results with the univariate analysis focused solely on pain-traits (figure *4.C*). Notably, we observed a significant increase in the mean top association statistic for 8 of the 13 K2P gene regions (KCNK3, KCNK4, KCNK5/16/17, KCNK6, KCNK7, KCNK9, KCNK10 and KCNK18) when incorporating both regular paracetamol use and pain in the multivariate model. The effect was particularly pronounced for the KCNK5 region, where the mean top association statistic (-Log P) for Pan-UK BB pain-related traits increased from 3.2 (univariate) to 5.3 (multivariate). Similar trends, albeit with more modest effects were also observed for other K2P regions when regular aspirin or ibuprofen use was considered in conjunction with pain (figure *4.C*).

These findings were further supported by local genetic correlation (Rg) analyses between regular analgesic use traits and each pain-related trait, for all the variants in each K2P gene region ±500 kb. These genetic correlations were consistent with the multivariate analysis results, whereby the largest proportion of local genetic correlations (rG >0.5) were between pain and regular paracetamol use (*supplementary figure 4*). Traits with the greatest No. correlations with regular analgesic use within K2P gene regions included multi-site chronic pain, back pain, tension headache, inflammatory neuropathy and painful respiration. Of note, the KCNK5 region exhibited a strong association with headache-related traits in which Rg >0.5 was observed between regular analgesic use and various headache phenotypes, including tension headache, other headache syndromes and migraine. Again, supporting previous evidence herein presented for this region and headache.

## Discussion

The complex and heterogeneous nature of chronic pain has historically limited the translational success of novel analgesic targets. Traditional genetic validation strategies have focused on exemplar gene families such as the voltage-gated sodium (SCNA) and transient receptor potential (TRP) channels, using small cohorts or rare familial mutations. Whilst such approaches have yielded key mechanistic insights, they have rarely translated into widely effective treatments. Broader genetic approaches, leveraging large-scale heterogeneous populations, may offer a more effective pathway for validating novel pain targets, especially for polygenic traits like pain.

Using GWAS summary statistics from over 500,000 individuals across UK and Finnish biobanks, we investigated the contribution of the two-pore domain potassium (K2P) family of ion-channels to pain-related phenotypes. Although these ion channels are known regulators of excitability and have been implicated in nociception through animal models (Pereira et al., 2014), expression data (Pollema-Mays et al., 2013, Gada & Plant, 2018) and small-scale human studies (Blanc et al., 2019, Langford et al., 2015 & Lafrenière et al., 2010), their relevance in large-scale human genetics has remained underexplored.

We highted pain-associated signals in 8 out of 13 K2P gene loci (±500 kb region), involving both rare and common (minor allele frequency >1%) variants spanning diverse pain phenotypes including neuropathic pain, headache, chest pain, and regular analgesic use. These findings position K2Ps as a potentially important gene family in human chronic pain.

To contextualise these findings, we applied identical analytical approaches to two benchmark ion channel families with established links to pain: SCNAs and TRPs. Significant single-variant associations were identified in four out of nine SCNA gene regions linked to headache and backpain, and multiple-variant associations were observed for gabapentin use and headache in three of the six TRPV regions. Strikingly, K2P gene loci performed comparably or in some cases, more robustly in terms of both statistical strength and breadth of phenotype association. These comparisons suggest that regulatory variants surrounding K2P genes may have broader relevance to pain susceptibility across populations than was previously appreciated. Furthermore, only one significant association was found between K2P genes and our randomly selected comparator traits (asthma, ischemic heart disease, peripheral nerve disorders and mental health disorders), indicating a degree of trait specificity for associations between K2P genes and pain phenotypes.

It is important to note that large scale GWAS analyses such as these may underrepresent the detection of rare, high-impact familial mutations. For example, a frameshift mutation in KCNK18 has been linked to migraine with aura in familial studies (Lafrenière et al., 2010) but showed no significant association in ours or other’s (Markel and Curtis, 2022) broader population-based analysis. Such discrepancies highlight the importance of scale-appropriate strategies, whereas rare variants may inform mechanistic understanding, common variant associations might offer translational value across more diverse populations.

Notably, most of the pain-linked variants identified in this study were located in non-coding regions more than 100 kb from K2P gene transcripts (*supplementary figure 3*). However, genomic distance from the gene of interest does not preclude functional relevance, as non-coding variants can exert long-range regulatory effects (Van Heyningen & Bickmore, 2013, Spitz, 2016) through mechanisms such as altered transcription factor binding, enhancer activity, or changes in chromatin accessibility. Functional validation of these variants, however, will be essential, particularly given that the ±500 kb region surrounding K2P genes may encompass other transcripts, complicating causal inference.

KCNK10 was the only K2P gene in this study with a Bonferroni-significant variant located within the gene transcript itself, which associated with pregabalin prescriptions, a treatment for chronic pain and anxiety. This adds weight to prior suggestions for the importance of KCNK10 in migraine (Royal et al., 2019). In contrast, while recent literature, primarily from animal studies, has highlighted KCNK2 as a promising analgesic target (Mathie et al 2021), we did not observe strong associations at this locus, emphasising the disparity that can exist between experimental and population-level data.

Of particular interest was the identification of a cluster of headache-associated variants (e.g. rs72854120 & rs72854118) in strong LD, within 50 kb of KCNK5, KCNK16 and KCNK17. These rare variants (minor allele frequency <1%) located within a promoter flank region were previously associated with headache in an Icelandic population (Bjornsdottir et al., 2023). A third variant, rs72851880, located just 14 kb from KCNK5 and residing within a CTCF transcriptional repressor site, was suggestively associated with headache, potentially exerting a minor protective effect. Downstream, (10-20 kb from KCNK5) additional variants (e.g., rs9470992, rs10947785, rs2561412, and rs471158) showed suggestive associations with regular paracetamol use, further strengthened in multivariate models. In support of this, a FinnGen variant (rs228822), located in the same genomic region, was significantly associated with femoral nerve lesion and shared weak LD with a variant previously linked to migraine and diastolic blood pressure. Together, these findings highlight a potential regulatory hotspot implicated in pain near to KCNK5 worth further functional investigation.

Given the polygenic and highly heterogeneous nature of pain, it is particularly striking that several pain-associated K2P gene variants were in moderate to strong LD with SNPs previously linked to other traits including depression and inflammatory bowel disease (KCNK7), fasting plasma glucose and migraine (KCNK5), white blood cell counts (KCNK3), pulmonary artery enlargement (KCNK10) and circulating phylloquinone (KCNK9). Weaker LD links were also noted for neuroticism and anxiety (KCNK18), platelet count (KCNK15), cortisone levels (KCNK12), bipolar disorder and Crohn’s disease (KCNK10) (*supplementary table 7 & 8*). This trait overlap is perhaps unsurprising given the known roles of K2P ion channels in processes beyond nociception, including cardiac physiology, immune function and neurobiology (e.g., depression and anxiety).

Importantly, multivariate analysis that incorporated both pain phenotypes with regular analgesic use, revealed enhanced genetic signals at several K2P loci (including KCNK10 and KCNK5), compared to univariate analysis of pain alone. Regular analgesic use, defined as self-reported use of ibuprofen, aspirin or paracetamol on most days over the past 4 weeks, was used as a proxy for chronic medication reliance and assumes that shared genetic architecture may exist between pain susceptibility and treatment behaviour. This was most evident for regular paracetamol use, which yielded strong associations at the KCNK5 gene locus, further implicating it as a target of interest.

### Limitations

The data presented herein, offer important insights into the importance of K2P channels in pain in humans. However, without direct functional validation, the assignment of causality to non-coding regulatory variants remains inherently challenging. Many of the pain-associated variants identified in this study are located more than 100 kb from K2P genes, often in regions encompassing multiple genes, making it difficult to definitively attribute their effects to K2P loci. Furthermore, pain phenotyping in large biobank cohorts, frequently relies on self-reported data or prescription records, which may lack clinical granularity.

### Conclusion

Despite these challenges, these findings provide large-scale genetic evidence to associate variants in K2P gene regions with chronic pain phenotypes in humans and identify these channels as a novel underexplored family of analgesic targets. These data are consistent with previous evidence, primarily from animal models, supporting a role of K2P channels in nociception (Mathie & Veale 2015). The evidence for KCNK5 is particularly compelling with convergent support from rare variant associations, strong linkage disequilibrium clustering and multivariate signals and highlights this particular K2P channel as a novel potential therapeutic target for chronic pain. The human genetic evidence presented here underscores the value of large-scale, population-based genomics in uncovering new targets for complex, polygenic conditions like chronic pain.

## Methods

### Analysis of Pan-UK biobank and FinnGen GWAS summary statistics

To probe the association of K2P gene region variants with pain, summary statistics were accessed from the Pan-UK biobank (Pan-UK biobank team., 2020, Karczewski et al., 2025) and FinnGen release 5 portals (Kurki et al., 2023). These data were generated using SAIGE (Zhou et al., 2018), a mixed model logistic regression framework applied to 29,865,259 v3 UK biobank variants and 16,962,032 release 5 FinnGen variants. Variants located within ±500 kilobases (kb) of the 15 K2P gene transcripts, were retained for further analysis. To benchmark the strength of association, we compared these to variants within ±500 kb of transient receptor potential (TRPV) and voltage gated sodium channel (SCNA) genes, which are established targets for pain. The strength of variant-trait associations was determined using three levels of significance: global (P < 5×10^-8^, -log p-value 7.3), Bonferroni (P < 5×10^-7^, -log p-value 6.3) and suggestive (P < 5×10^-6^, -log p-value 5.3). The Bonferroni-corrected significance threshold was calculated based on a nominal alpha level (P = 0.05) divided by the maximum number of independent K2P gene variants tested for. Additionally, a suggestive significance threshold was set to identify variants of potential, where at least one false positive per genome-wide scan is expected (Cheverud, 2001).

Variants were mapped to the GRCh37 or GRCh38 genome builds in relation to gene and enhancer/promoter positions using the genome UCSC table browser (Karolchik et al., 2004). Mapped variants were visualised using GraphPad PRISM 9.5.0. RS ID numbers were assigned, and variants annotated using SNPNEXUS (Dayem Ullah et al., 2018). For variants of interest, functionality was assessed using a suite of computational tools designed to predict the likelihood of nucleotide base changes affecting gene regulation and/or expression. Specifically, we used the following tools: RegulomeDB (Dong et al., 2023), Combined Annotation Dependent Depletion (CADD) (Rentzsch et al., 2021). Fitness Consequences of Functional Annotation (fitCons) (Gulko et al., 2015), Eigen, an unsupervised spectral approach for scoring variants (Ionita-Laza et al., 2016), Functional Analysis through Hidden Markov Models (FATHMM) (Shihab et al., 2012), FunSeq2 (Fu et al., 2014) and Regulatory Mendelian Mutation (ReMM) (Smedley et al., 2016). Previously reported trait associations were identified using the GWAS Catalog (Cerezo et al., 2024). LDtrait (Lin et al., 2020) and LDlink together with FORGE annotation (Machiela & Chanock, 2015) were used to identify linkage disequilibrium (LD) between variants of interest and previously reported trait-associated variants, as well as to generate LD plots for visualisation. LD is defined as the non-random association (co-inheritance) of alleles (variants) on chromosomes.

### Selection of pain-related and comparator traits

A total of 136 traits (*supplementary table 1 & 2*) were extracted for analysis from the Pan-UK BB (Pan-UK biobank team., 2020, Karczewski et al., 2025) (https://pan.ukbb.broadinstitute.org/downloads) and FinnGen phenotype manifests (Kurki et al., 2023) (https://r5.risteys.finngen.fi/). These included pain-related conditions (migraine, fibromyalgia, neuropathies), pain categorised by body region (head, back, neck) and medication usage (opioid, non-steroidal anti-inflammatory, antipyretic). Where possible, data was categorized as acute or chronic (defined as persisting for more than 3 months). Traits with fewer than 3000 cases were considered as having low statistical power. For the Pan-UK BB dataset, phenotypic data included a combination of self-reported responses collected via questionnaires or nurse-led interviews, as well as primary care and electronic health records. In contrast. the FinnGen dataset was derived exclusively from health registry data recorded by healthcare professionals. To test the robustness of this approach, we included data from 3 randomly selected, well powered, Pan-UK BB traits with >10,000 cases as comparators.

### Multivariate analysis of GWAS summary statistics to enhance trait-associated loci discovery

To increase the detection of trait-associated loci and further explore the pain-linked effects of variants we applied MultiABEL (Shen et al., 2017, Ning et al., 2021), a multivariate method which uses GWAS summary statistics to implement a Pillai’s trace MANOVA of multiple phenotypes against each variant. For this, we generated a multivariate p-value for all common K2P gene variants (allele frequency > 1%) across all non-medication pain-related traits from the Pan-UK BB, in combination with one of three chronic analgesic-use traits: regular use of aspirin, paracetamol, or ibuprofen. In this way we aimed to capture variants associated with both pain phenotypes and chronic analgesic use. The same approach was used to collate local genetic correlation scores for paired pain and analgesic traits within K2P gene regions as a means of identifying shared genetic underpinnings. Multivariate affects were considered strong when the multivariate association resulted in at least an order of magnitude improvement in the P-value compared to the corresponding univariate association for the same pain trait.

## Data Availability

All data produced in the present study are available upon reasonable request to the authors

https://pan.ukbb.broadinstitute.org/downloads

https://finngen.gitbook.io/documentation/data-download

## Author contributions

The study was conceived and coordinated by S.R.B, E.L.V, A.M, P.D.W, J.J and R.F. Bioinformatic analysis and data acquisition was performed by S.R.B and technically advised by J.C. Interpretation of results was performed by S.R.B, E.L.V and A.M. Writing of the paper was performed by S.R.B, E.L.V and A.M. Edits and final approvals of paper were performed by S.R.B, E.L.V, A.M, P.D.W, J.J, J.C and R.F. All authors agree to be held accountable for and approve of the content within this manuscript.

## Data availability

Source GWAS summary statistics supporting this study are available from the Neale Lab UKB round 2 GWAS (http://www.nealelab.is/uk-biobank/ukbround2announcement) and FinnGen release 5 (https://finngen.gitbook.io/documentation/r5/data-download). Processed and filtered data is available within the supplementary files contained within this manuscript. Raw data from the multivariate analysis is available upon request.

## Code availability

The open source R package and tutorial for MultiABEL is available at https://cran.r-project.org/package=MultiABEL and https://github.com/xiashen/MultiABEL/. Code used for the generation of pan-UK biobank summary statistics from the Neale Lab UKB GWAS round 2 is available at https://github.com/atgu/ukbb_pan_ancestry and at https://github.com/FINNGEN/ for the analysis of FinnGen GWAS results.

## Acknowledgments

We want to acknowledge the participants and investigators of the FinnGen study, UK Biobank and Pan-UK Biobank study. The Pan-UK Biobank study was originally conducted under project ID 31063. A.M and E.L.V were supported by a LifeArc Centre for Therapeutics Discovery Award. S.R.B was supported by an industrial fellowship award from the Royal Commission for the Exhibition of 1851.

## Materials & Correspondence

Please address correspondence and material requests to S.R.B

---

**Supplementary figure 1.**
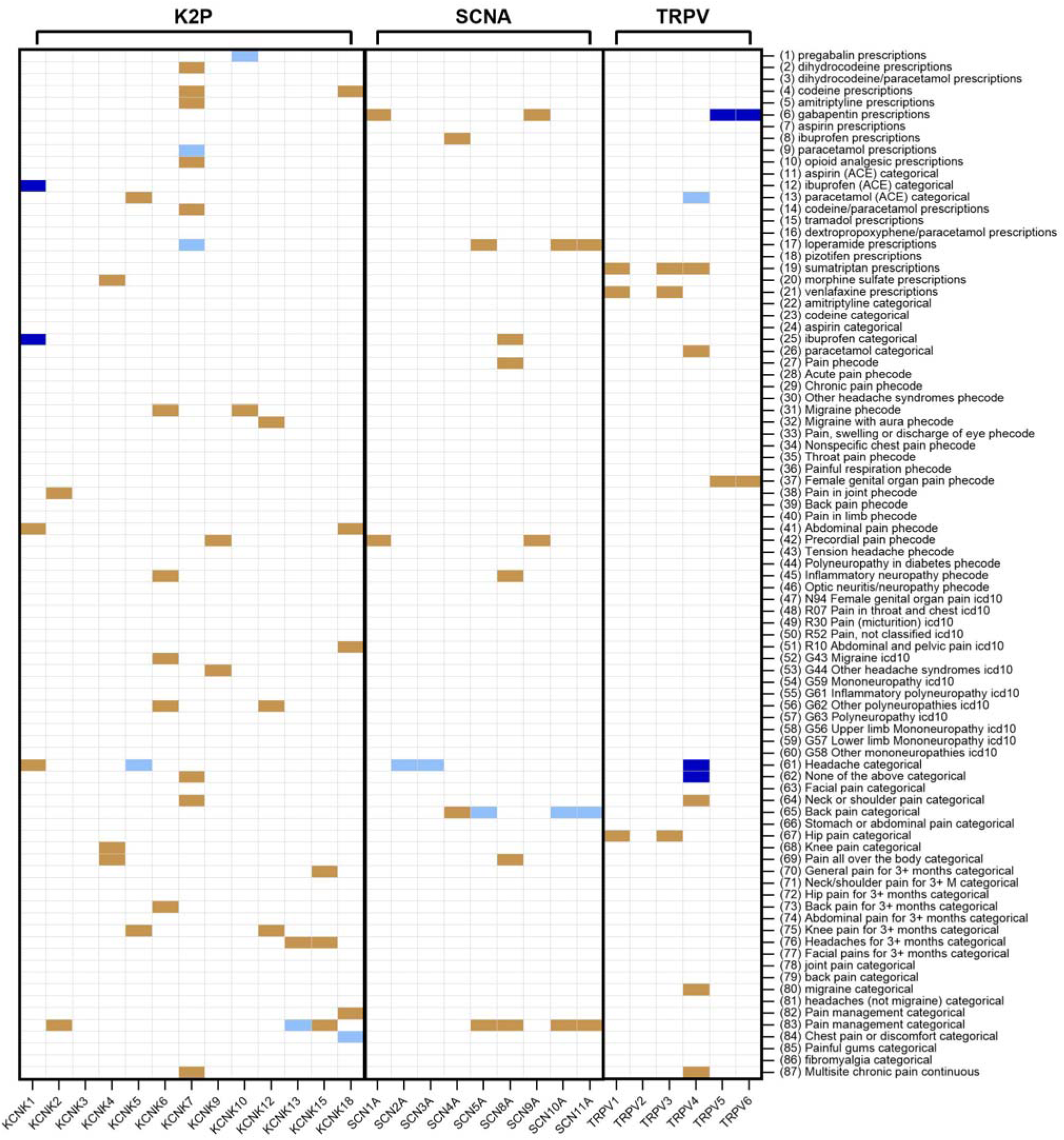
Analysis of the distribution and specificity of Pan-UK biobank pain-associated variants in K2P, SCNA and TRPV gene regions. Each cell represents the maximum - log p-value for the association of gene variants within 500 kb of each gene transcript, with each of the 87 pain-related phenotypes. Trait numbers in brackets correspond to those in supplementary table 3. Medication traits refer to either prescription data or self-reported regular use. Significant - log p-value associations are shown for suggestive (-log p-value > 5.3 – brown), Bonferroni (-log p-value > 6.3 – light blue) and global (-log p-value > 7.3 – dark blue) significance levels.

**Supplementary figure 2.**
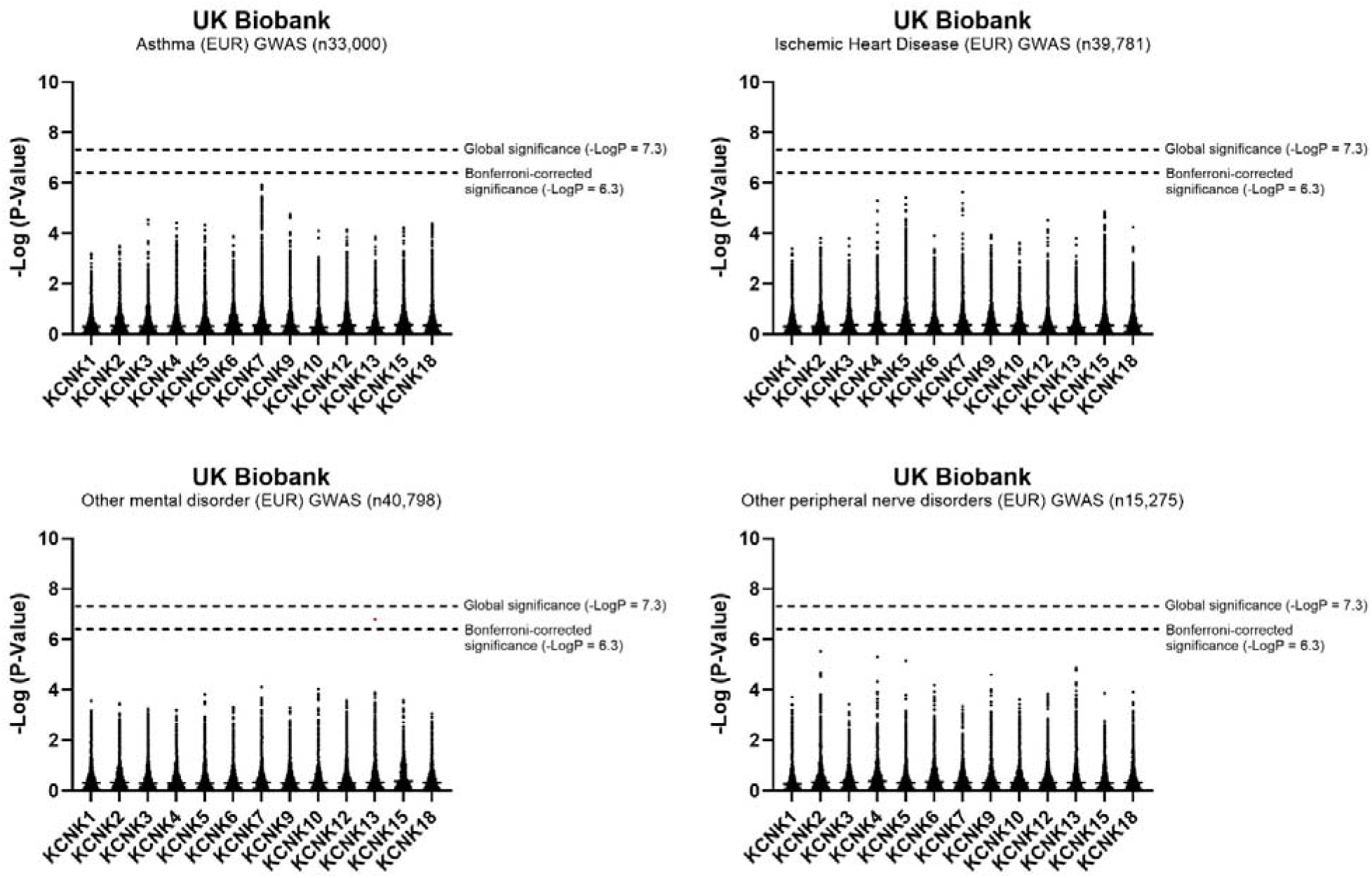
Pan-UK Biobank genome-wide association summary statistics from 3 randomly selected ‘phecode’ traits with >10,000 cases, for the association of gene variants within 500 kb of K2P gene transcripts.

**Supplementary figure 3.**
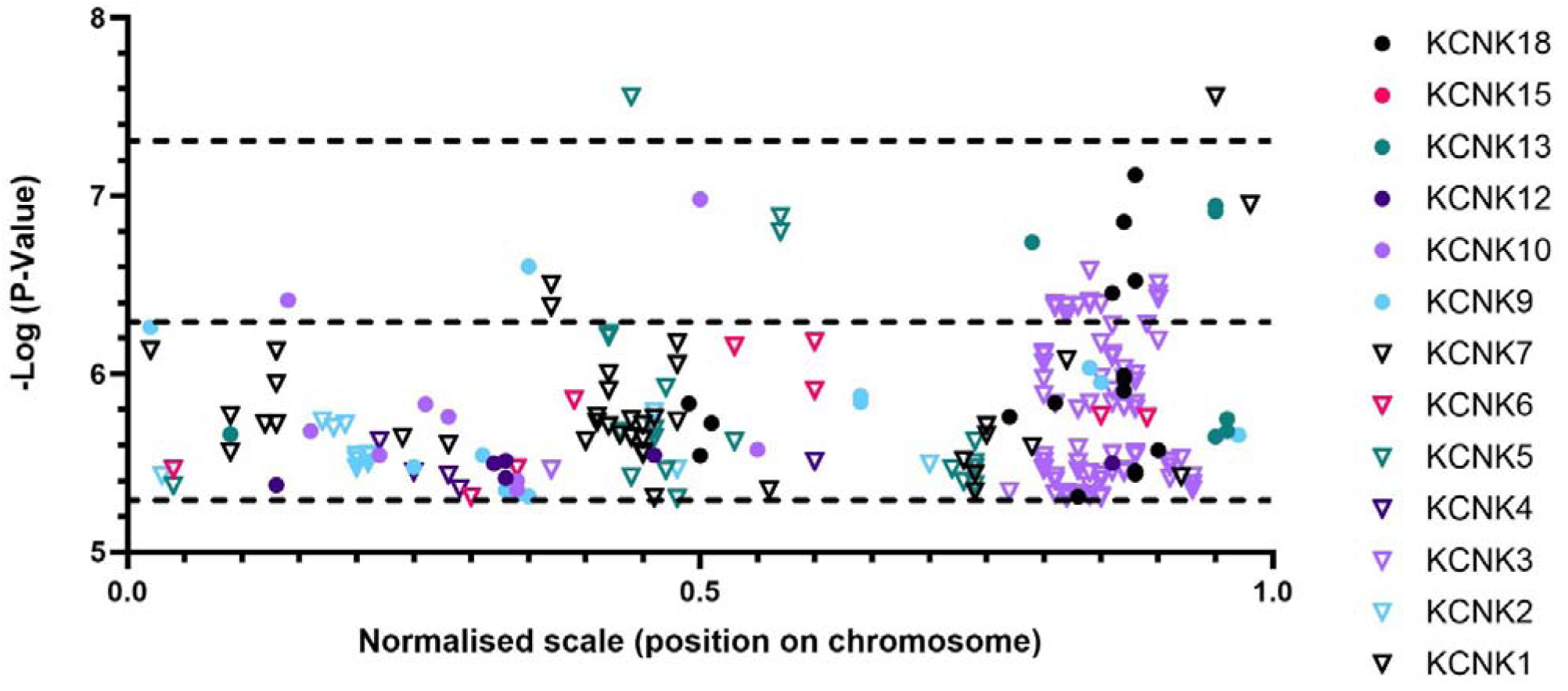
Spatial mapping of variants with Bonferroni (-log P > 6.3) and suggestive (-log P > 5.3) significant level associations with pain in K2P gene loci (+/- 500KB) on a normalized scale. With 0 representing –500 KB, 0.5 the centre of the K2P transcript, and 1 representing +500 KB.

**Supplementary figure 4.**
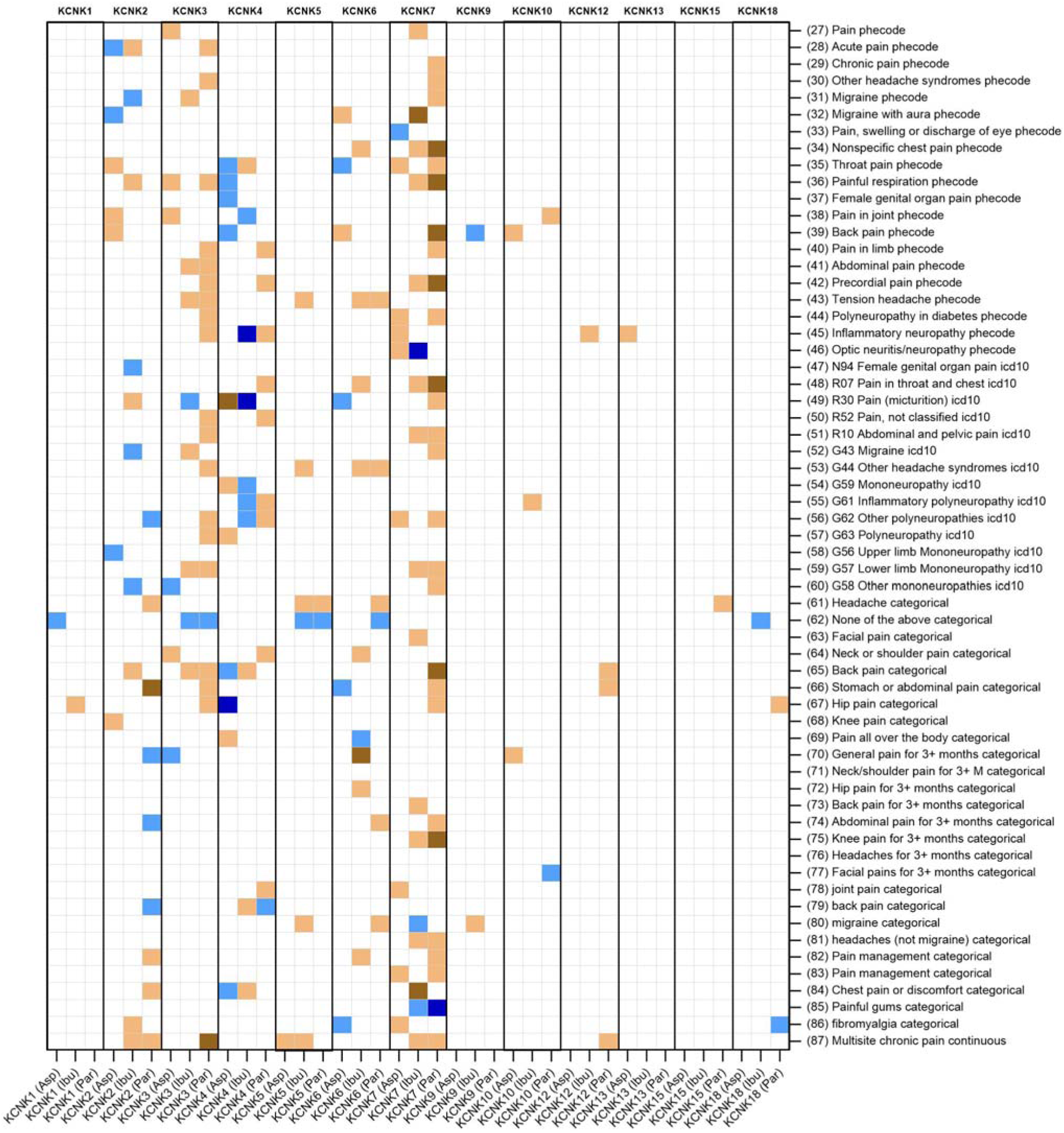
Local genetic correlations (rG) of all gene variants within 500 kb of K2P transcripts, between Pan-UK biobank non-medication pain-related traits and regular analgesic medication use (aspirin (asp), paracetamol (par) or ibuprofen (ibu)). Trait numbers in brackets correspond to those in supplementary table 3. rG was categorized as positively correlated between 1 and 0.75 (dark brown), moderate between 0.75 and 0.5 (light brown), negatively between -1 and -0.75 (dark blue) and moderately negative between -0.75 and -0.5 (light blue).

